# Association of Travel Burden with Definitive Prostate Cancer Treatment: A United States Registry Cohort Study

**DOI:** 10.1101/2025.09.26.25336503

**Authors:** Filippo Dagnino, Stephan Korn, Danesha Daniels, Zhiyu Qian, Daniel Stelzl, Hanna Zurl, Klara Pohl, Mei-Chin Hsieh, Brenda Y. Hernandez, Andrea Piccolini, Giovanni Lughezzani, Nicolò M. Buffi, Stuart R. Lipsitz, Amanda Reich, Joel S. Weissman, Alexander P. Cole, Quoc-Dien Trinh, Hari S. Iyer

## Abstract

**Purpose:** Prostate cancer (PCa) mortality disparities are partly driven by unequal access to care. Transportation barriers may limit access to definitive treatment. We studied how driving travel time affects receipt of definitive PCa treatment.

**Materials and Methods:** We conducted a cohort study of men with non-metastatic PCa (2000 - 2015; follow-up through 2018) across the metropolitan area cancer registries of seven US states. Travel burden was estimated using Google Maps isochrones representing driving time thresholds to reach the hospital appended to geomasked residential addresses. Outcomes were “no treatment, “ “radical surgery,” or “radiotherapy”. Covariate-adjusted multinomial logistic regression with interaction terms assessed modification by sociodemographic factors.

**Results:** The study included 132,939 men, of whom 37.0% received no treatment, 41.0% underwent surgery, and 22.0% received radiotherapy. Longer driving time (≥90 min vs <30 min) was associated with higher radical prostatectomy (aOR: 1.07, 95% CI: 1.03, 1.12), but lower radiotherapy (0.72, 95% CI: 0.69 - 0.76). Subgroup analyses revealed higher surgery associated with longer driving times among those in nSES Q1 (aOR: 1.33, 95% CI: 1.21-1.45) vs Q5 (aOR: 0.94, 95% CI: 0.86-1.04), those in low (aOR: 1.16, 95% CI: 1.09-1.24) vs high (aOR: 1.03, 95% CI: 0.98-1.09) population density areas, and those with regional (aOR: 1.30, 95% CI: 1.14-1.48) vs localized (aOR: 1.05, 95% CI: 1.00 -1.09) disease. Longer driving time was mostly associated with lower odds of radiotherapy across sociodemographic subgroups.

**Conclusions:** Higher travel burden was associated with lower radiotherapy receipt, but greater surgery use in deprived and rural patients, which warrants further investigation.

## INTRODUCTION

Prostate cancer (PCa) is the most prevalent non-cutaneous cancer among men in the US, with 313,780 new cases expected in 2025 ^1^. Previous research shows that PCa poses a greater mortality risk for Black men compared to White men ^2,3^, with the disparity in PCa-related deaths between these groups widening over recent decades ^4,5^. This growing gap may, in part, be attributed to persistent racial disparities in accessing prompt and effective treatment ^2^. Recent evidence indicated that Black men are more likely than White men to receive definitive PCa treatment at under-resourced facilities ^6,7,8^. Moreover, care received at institutions serving predominantly marginalized populations, such as Black communities, are associated with less favorable treatment outcomes ^9^.

Access to care is a multifaceted concept, predominantly affected by financial determinants such as insurance coverage ^10,11^. However, various non-financial factors can also influence healthcare accessibility. Notably, limited evidence exists regarding the impact of travel burden on accessing equitable PCa care. Travel burden is defined as the time required to travel from a patient’s residential address to the facility where PCa care is provided ^12^.

Evidence from the literature indicates that Black patients typically face greater travel burden than White patients when seeking care, as they are more likely to live farther from healthcare facilities, requiring longer journeys to access treatment ^13-17^. This disparity can heighten barriers to healthcare, leading to delays in treatment, lower treatment rates, and poorer outcomes ^18^. While the influence of travel burden on the management of chronic medical conditions is well-documented ^14,19,20^, its impact on cancer care, particularly PCa care, remains underexplored. Understanding how travel burden can exacerbate racial disparities in access to definitive PCa treatment is crucial to identify strategies to enhance timely treatment and mitigate inequities in outcomes across racial groups. This study examined the relationship between the travel time spent driving from a patient’s residence to the PCa treatment facility and the receipt of definitive PCa treatment, and whether this association varied by sociodemographic factors.

## METHODS

### Data Source and Study Population

Using data from the Multilevel Epidemiologic Tumor Registry for Oncology (METRO)—a registry database of men with PCa from multiple US states - we constructed a cohort of men aged 40 years and older diagnosed with PCa between 2000 and 2015 from Hawaii, Louisiana, Massachusetts, New Jersey, Ohio, Utah, and Washington (Seattle/Puget Sound metropolitan area) (n=423,210) ^21^. These states were selected based on availability of reporting facility information to calculate residential travel time to facility where patients were diagnosed or treated, and to represent the four U.S. Census regions (Northeast, Midwest, South, and West). Patient follow-up began at diagnosis and continued through the end of 2018. Geomasked residential address geocodes at the time of diagnosis were obtained ^22^. Data included address-level information on residential neighborhood context and the facilities where PCa treatment was reported.

We excluded participants who were <40 or ≥99 years old (n=306), those who were diagnosed at autopsy (n=10,045), and those for whom reporting facility and residential address were unavailable (n=252,474). We further excluded participants with SEER summary stage of “distant” or “missing” because these were unlikely to be eligible for definitive treatment (n=27,446), for a final analytic sample (n=132,939).

### Outcome

The primary outcome was defined as the timely receipt of definitive PCa treatment, categorized as undergoing either radical surgery or radiotherapy within 90 days of diagnosis, or receiving no definitive treatment. Cancer registries capture the initial course of treatment (within four months after diagnosis) for surgery or radiation therapy, and the date when this event took place. Trained registrars review patient medical records and update information as it becomes available ^23^. While no formal guidelines specify exact timeframes for timely PCa treatment, starting therapy within 90 days is generally deemed sufficient to accommodate logistical challenges and potential delays ^24^.

### Predictor

Travel burden, defined as the driving travel time required for a patient to travel from residence to the hospital where treatment was received, was the primary predictor of the analysis. We integrated detailed estimates of patients’ residential addresses, facility locations, and road routes used to access care with registry data on the treatment facility to model actual, rather than potential, access. To minimize computational demands, we created travel time zones around each facility where PCa patients sought care. Therefore, isochrones were generated using Google Maps’ routing algorithm using the Travel Time API ^25^, which combines road network data with machine learning models to provide real-time driving travel time estimates. Isochrones delineated polygonal zones, indicating area boundaries around each facility where patients would have taken the longest time to reach care, based on different driving time thresholds (<30, 30–59, 60–89, and ≥90 minutes driving travel time). These thresholds were calculated through the Google Maps routing algorithm via the Travel Time API. These isochrones were linked to patients’ geomasked residential addresses, enabling the estimation of driving travel time to the facility where treatment was actually received.

### Covariates

The analysis included sociodemographic characteristics of patients including age at diagnosis, year of diagnosis, race and ethnicity (non-Hispanic White, non-Hispanic Black, Hispanic, Asian American, Native Hawaiian, Pacific Islander), nSES (using an index previously developed from a subsample of the included registries) ^4^, SEER summary stage based on available clinical and pathological data (Localized vs Regional) ^26^, health insurance status (private, uninsured, Medicaid, Medicare, Other government), and population density (<1000 people/mi^2^, ≥1000 people/mi^2^). Census tract-level socioeconomic data were sourced from the 2000 decennial Census and the 2010 American Community Survey and linked to participants’ geomasked residential address at time of diagnosis as described previously ^21,22^.

### Statistical Analysis

Proportions of different definitive treatment types, driving travel times, and sociodemographic characteristics of the cohort were summarized using descriptive statistics. A multinomial logistic regression model was used to calculate adjusted odds ratios (aOR) and 95% confidence intervals (95% CI) for driving time and other predictors of receiving timely “radical surgery” or “radiotherapy” versus “no treatment”. Additionally, we incorporated mulitiplicative interaction terms to assess how race, nSES, population density, and stage modified the likelihood of undergoing surgery or radiotherapy compared to no treatment, for each driving time threshold. Likelihood ratio tests were used to evaluate statistical significance of interactions. We used multiple imputation with chained equations for missing data on nSES, insurance, and population density using predictive mean matching for continuous variables and polytomous logistic regression for categorical variables using the mice package in R ^27^. All reported P-values were two sided and we considered a value of less than 0.05 to be significant. Data manipulation and analyses were performed using R version 4.3.0 (R Foundation, Vienna, Austria).

## RESULTS

### Baseline Characteristics

A total of 132,939 patients met the inclusion criteria. Participants who were excluded were more likely to be diagnosed from 2000-2004, reside in the lowest quintile of neighborhood socioeconomic status (nSES), reside in high population density areas, and less likely to report radical prostatectomy (**Supplementary Table 1**). The mean age was 65.8 years (SD=9.2). Most of the cohort identified as Non-Hispanic White (77.9%), followed by Non-Hispanic Black (12.9%). Among eligible patients, 37.0% did not receive treatment, 41.0% underwent radical surgery, and 22.0% received only radiotherapy. Most patients (43.1%) had a driving travel time <30 minutes to the hospital where PCa treatment was provided. Compared to men with PCa <30 minutes from the hospital where they sought care, those ≥90 minutes were more likely to be NHW (82.2% vs 73.1%), less likely to have Medicare insurance (24.4% vs 41.0%), less likely to have Gleason Score <7 (19.2% vs 25.7%), and more likely to reside in low population density areas (50.6% vs 22.8%). Additional participant characteristics of the cohort are reported in **Table 1**.

**Table 1.**
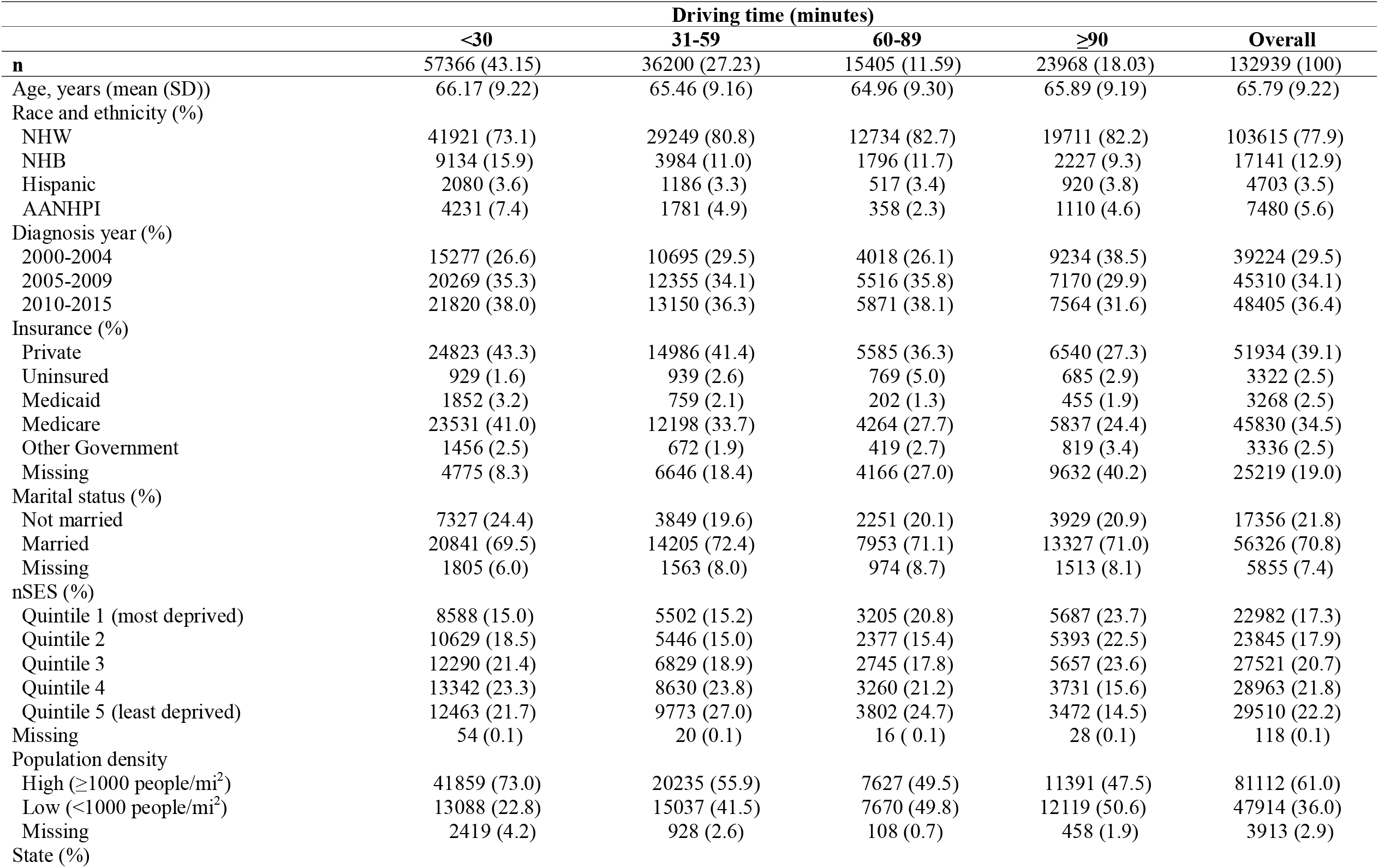
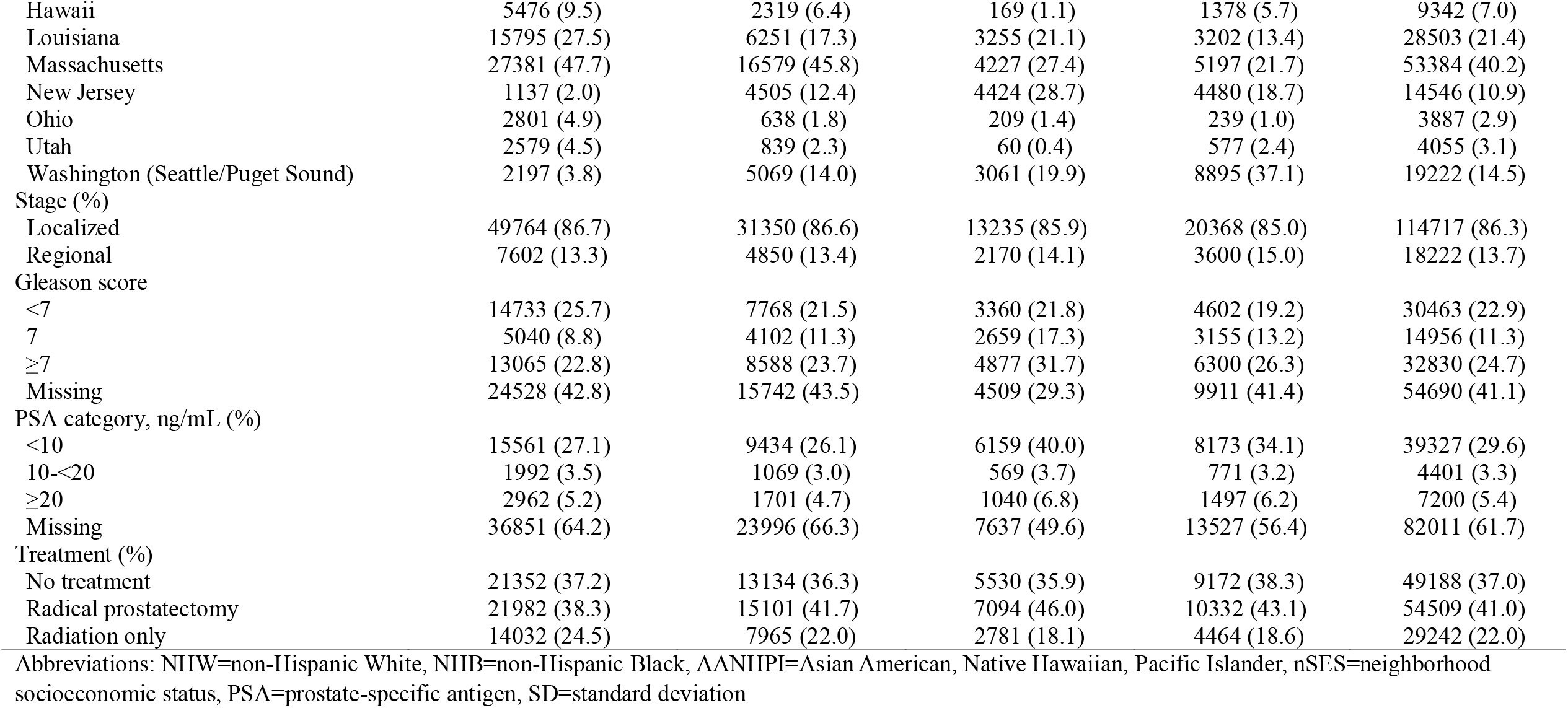
Descriptive characteristics of men with prostate cancer diagnosed with localized and regional stage.

### Association of driving time with surgery and radiotherapy compared to no treatment

Compared to individuals with <30 minutes driving time to treatment, those with times exceeding 30 minutes had higher odds of surgery compared to no treatment (31-59 minutes: OR: 1.12, 95% CI: 1.08-1.15, 60-89 minutes: OR: 1.25, 95% CI: 1.20-1.30, ≥90 minutes: OR: 1.09, 95% CI: 1.06-1.13; **Table 2**). Following covariate adjustment, compared to individuals with <30 minutes driving time to treatment, there was attenuation of estimates but longer travel time remained associated with higher odds of surgery compared to no treatment (31-59 minutes: OR: 1.01, 95% CI: 0.98-1.05, 60-89 minutes: OR: 1.13, 95% CI: 1.08-1.18, ≥90 minutes: OR: 1.07, 95% CI: 1.03-1.12).

**Table 2.**
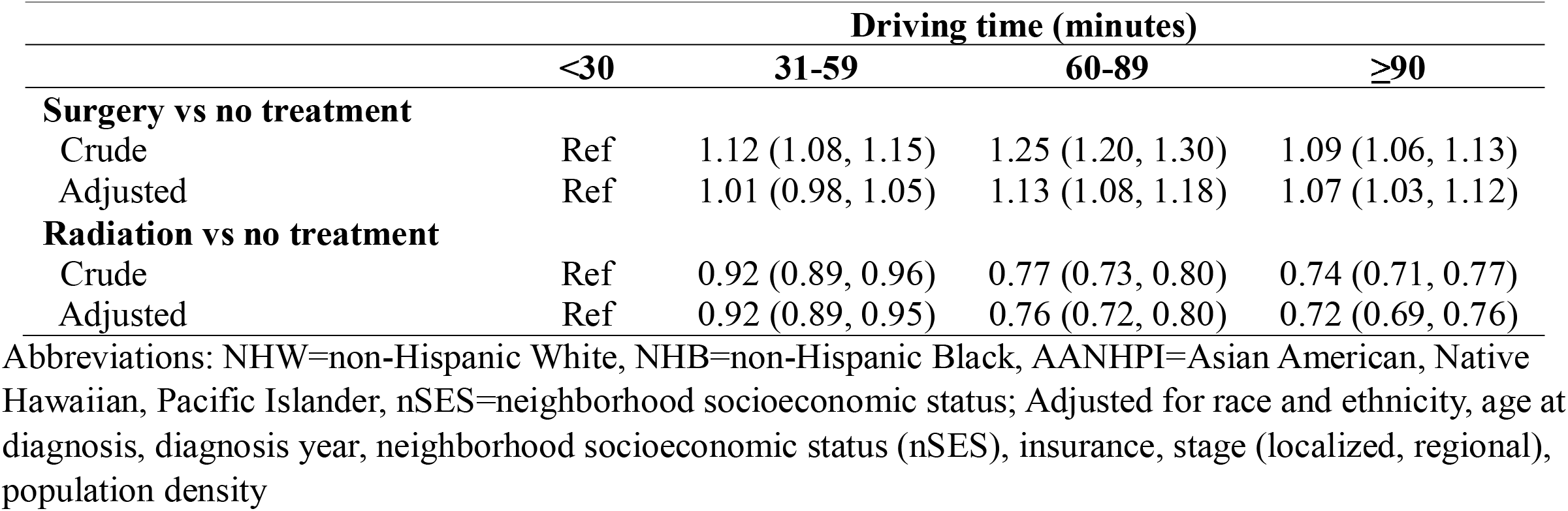
Association of driving travel time from residence to treating facility from multinomial regression models for surgery and radiotherapy vs no treatment.

Longer driving time was associated with lower odds of receiving only radiotherapy compared to no treatment (**Table 2**). From fully adjusted models, compared to individuals with <30 minutes driving time to treatment, those with times exceeding 30 minutes had lower odds of radiotherapy compared to no treatment (31-59 minutes: aOR: 0.92, 95% CI: 0.89-0.95, 60-89 minutes: aOR: 0.76, 95% CI: 0.72-0.80, ≥90 minutes: aOR: 0.72, 95% CI: 0.69-0.76).

### Stratified associations of driving time with surgery and radiotherapy

We examined whether associations of driving time with receipt of surgery and radiotherapy varied by race and ethnicity, nSES, population density, and stage (**Table 3**). Associations of driving time with receipt of surgery vs no treatment varied by race (*P*_het_ <.001). Compared to individuals with <30 minutes driving time, those with ≥90 minutes driving time had similar odds of surgery for NHW (aOR: 1.07, 95% CI: 1.03-1.12) and AANHPI (aOR: 1.01, 95% CI: 0.85-1.20), but not NHB (aOR: 1.14, 95% CI: 1.01-1.28) or Hispanic individuals (aOR: 1.17, 95% CI: 0.96-1.43). Similarly, associations of driving time with receipt of surgery vs no treatment varied by nSES (*P*_het_ <.001).

**Table 3.**
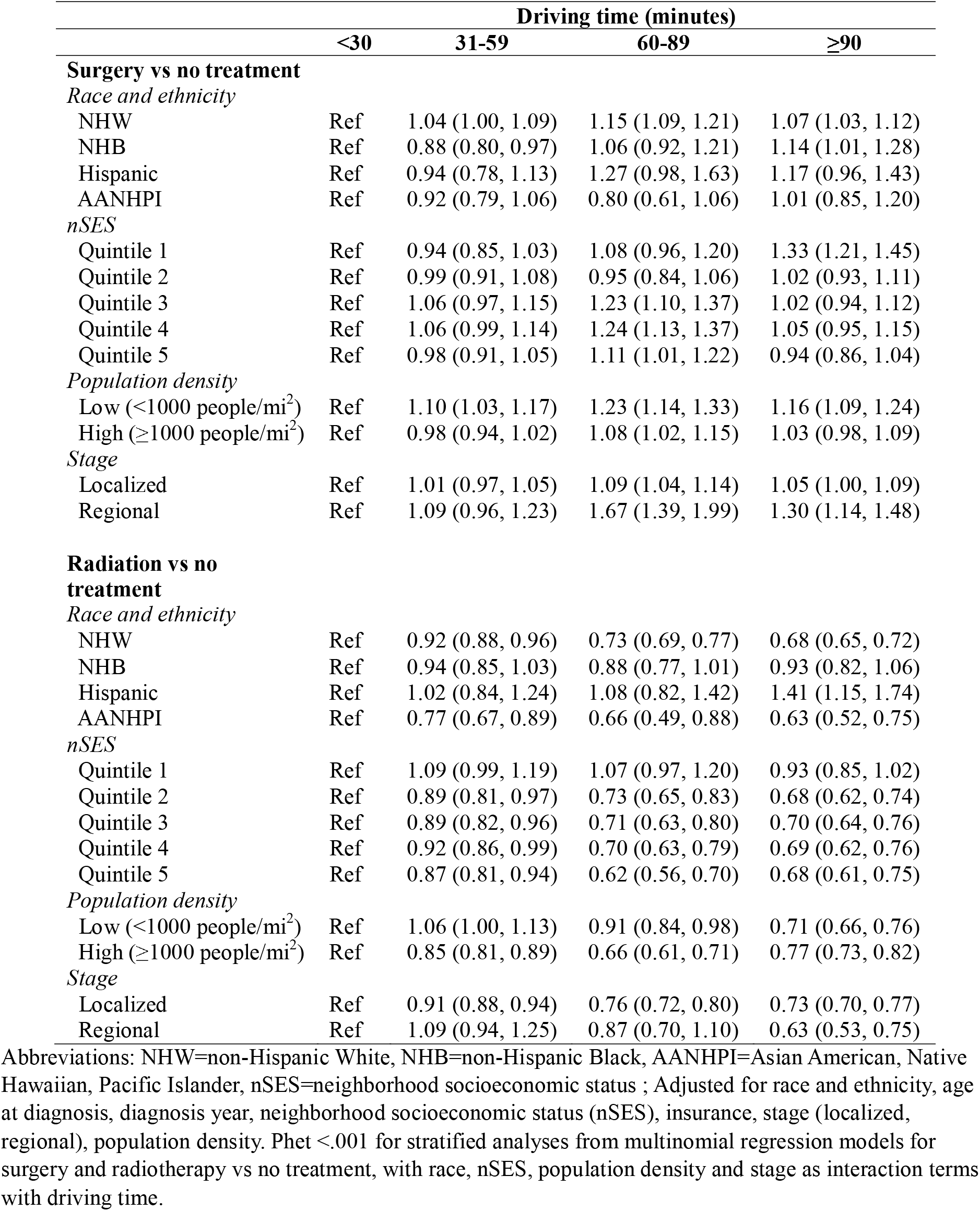
Association of driving travel time from residence to treating facility from multinomial regression models for surgery and radiotherapy vs no treatment, stratified by race and ethnicity, neighborhood socioeconomic status, population density, and stage.

Longer driving time was associated with higher odds of surgery compared to no treatment for those in nSES quintile 1 (most deprived) (aOR ≥90 vs <30 minutes: 1.33, 95% CI: 1.1-1.45), but was associated with not statistically significant lower odds of surgery among those in nSES quintile 5 (least deprived) (aOR ≥90 vs <30 minutes: 0.94, 95% CI: 0.86-1.04). Longer driving time was associated with higher odds of surgery among individuals in low (aOR: 1.16, 95% CI: 1.09-1.24) but not high population density areas (aOR: 1.03, 95% CI: 0.98, 1.09); and among both those with regional (aOR: 1.30, 95% CI: 1.14-1.48) and localized stage (aOR: 1.05, 95% CI: 1.00-1.09).

For associations of driving time with odds of receiving only radiotherapy, there was significant variation across subgroups of race, nSES, population density, and stage. Longer driving time was associated with lower odds of receiving radiotherapy for NHW (aOR ≥90 vs <30 minutes: 0.68, 95% CI: 0.65-0.72) and AANHPI (aOR: 0.63, 95% CI: 0.52-0.75), but this was not the case among NHB (aOR: 0.93, 95% CI: 0.82-1.06) and Hispanic (aOR: 1.41, 95% CI: 1.15-1.74) individuals. For those in the lowest quintile of nSES (most deprived), there was higher odds of receipt of radiotherapy among those with 31-59 minutes (aOR: 1.09, 95% CI: 0.99-1.19) and 60-89 minutes (aOR: 1.07, 95% CI: 0.97-1.19) but not ≥90 minutes (aOR: 0.93, 95% CI: 0.85-1.02) compared to those with <30 minutes of driving time, though these did not reach statistical significance. However, higher driving time was associated with lower receipt of radiotherapy for individuals in increasing quintiles of nSES. Associations of longer driving time with lower receipt of radiotherapy were weaker among individuals with low compared to high population density, and among those with localized compared to distant stage.

## DISCUSSION

In this study, we evaluated how the burden of driving travel time influenced the receipt of definitive PCa treatment and whether associations varied by race, nSES, population density, and stage. Our findings suggested that longer travel times were associated with a decreased likelihood of receiving only radiotherapy compared to no treatment but were not associated with lower receipt of surgery overall. The primary multinomial model revealed differing outcomes for surgery and radiotherapy receipt based on race, nSES, population density, and stage.

Despite the wide confidence intervals, increased odds of surgery receipt were reported with longer driving time in Hispanic individuals. We found that longer driving time was associated with higher odds of surgery among individuals in the most deprived quintile of nSES but not in the other quintiles, which may partly reflect urban-rural differences in travel time and nSES ^20^. Similarly, longer driving time was associated with higher odds of surgery among those in rural but not urban areas. In addition, longer driving time was associated with higher odds of receipt of surgery among those with localized and, even more so, regional stage disease, which could imply that those farther from care may delay treatment, leading to more advanced disease requiring intervention when they do eventually seek care.

Conversely, longer driving time was associated with lower odds of receiving radiotherapy among NHW and AANHPI patients, while this association was less clear for NHB and Hispanic individuals. Among patients in the most disadvantaged nSES quintile, driving 31–59 or 60–89 minutes was associated with higher odds of radiotherapy receipt, although this trend did not persist for longer travel times. In contrast, among patients in higher nSES quintiles, longer driving times were associated with lower radiotherapy receipt. The negative impact of longer travel time on radiotherapy receipt was also weaker among those living in sparsely populated areas compared to densely populated ones, and among patients with localized versus distant-stage PCa. These differences may reflect variations in clinical presentation and treatment practices between urban and rural facilities, which should be further explored.

Previous studies suggest that both urban and rural patients are less likely to receive radiotherapy compared to radical surgery as the distance from a treatment facility increases ^28^. The centralization of robot-assisted radical prostatectomy (RARP) in high-volume centers—where perioperative outcomes have significantly improved ^20,29^—may encourage patients to travel long distances to access these specialized facilities. This pattern has also been observed in the surgical management of other cancer types ^30^. On the other hand, radiotherapy for PCa requires daily sessions over several weeks, placing a considerable travel burden on eligible patients. As a result, many individuals facing substantial travel challenges may opt for a single long-distance trip to undergo radical surgery at a high-volume center rather than repeatedly commuting for radiotherapy sessions. Since socioeconomic factors also impact travel burden, travel time may serve as an additional indicator acting as a proxy variable to highlight disparities in access to appropriate PCa treatment ^20^.

Our study is among the first to leverage a geospatial routing algorithm based on Google Maps that integrates road network data with machine learning models to provide real-time travel predictions. We leveraged data from a large, geographically representative population-based cohort for radical treatment receipt of localized PCa, encompassing a broad range of socioeconomic backgrounds and racial groups. Additionally, the study population spans all four US regions (West, Midwest, Northeast, and South), providing a comprehensive perspective on the relationship between travel burden and access to PCa care across the entire country. This study could yield information regarding travel patterns in relation to treatment receipt which can be used to develop strategies for reducing travel-related challenges in accessing PCa care.

### Limitations

We were unable to capture the exact facility location where participants received care due to privacy restrictions imposed by registries. We used the reporting facility as a proxy for the facility where patients actually sought care, which may introduce misclassification bias. However, this is likely to be non-differential and therefore attenuate results towards the null. Second, because this study only assesses completed treatment visits, we are unable to evaluate whether surgery or radiotherapy was offered but patients chose not to pursue it due to travel or other access barriers, a challenge in all administrative databases. However, evaluating associations of driving time with receipt of different treatments across sociodemographic subgroups can provide guidance for populations who may face greater travel barriers when seeking care (rural patients, patients with low nSES). Lastly, we were unable to capture reporting facilities for all participants, and if this missing pattern was correlated with treatment, selection bias may distort associations. However, our analytic sample still reflects diversity with respect to race and ethnicity, nSES, insurance, and states across the US.

## CONCLUSIONS

We found that longer driving times were associated with lower receipt of only radiotherapy, but not with surgery, compared to no treatment in a population-based sample of men with PCa from multiple US states. Subgroup analyses revealed that men residing in low nSES neighborhoods and those from rural areas had higher odds of surgery associated with longer driving times, suggesting opportunities for outreach and social supports to reduce travel burden among those in need of surgery in these groups.

## Data Availability

The data that support the findings of this study are available from the corresponding author upon reasonable request. Data for this study were requested from each registry for the purposes of this research, and Institutional Review Board and Data Use Agreements between Rutgers, Dana-Farber, and each registry prohibit sharing of these data outside of the research team. Inquiries can be directed to

## ACKNOWLEDGMENTS

**QDT** reports consulting fees from Astellas, Bayer, Intuitive Surgical, Janssen, Novartis, Pfizer, and research funding from the American Cancer Society, Pfizer Global Medical Grants (Prostate Cancer Disparities #63354905), and a Health Disparity Research Award from the Department of Defense Congressionally Directed Medical Research Program (#PC220551).

**APC** reports research funding from the Bruce A Beal and Robert L Beal surgical fellowship of the BWH Department of Surgery, from the Prostate Cancer Foundation and American Cancer Society (#23YOUN25) and from a Physician Research Award from the Department of Defense Congressionally Directed Medical Research Program (#PC220342). The other co-authors declare that they have no known competing financial interests or personal relationships that could have appeared to influence the work reported in this paper.

**HIS** reports research funding from Prostate Cancer Research, UK and the National Institutes of Health (K01ES035734).

